# Quantifying the Trainability of Peripheral Nerve Function in Young and Older Adults

**DOI:** 10.1101/2024.10.01.24314722

**Authors:** JoCarol E. Shields, Claire M. Smith, Shawn M. Reese, Marcel L. Dos Santos, Maria Parodi, Jason M. DeFreitas

## Abstract

It is well known that the natural progression of age can result in motor neuron degeneration. Consequently, this leads to slowing of nerve conduction, denervation, and reduced motor function. Slower neural conduction can negatively alter an individual’s response time, which could increase the risk of falls. Further investigation is needed to determine the potential role exercise interventions may afford in mitigating age-related nerve deterioration. The purpose was two-fold: first, to determine the effects of resistance training on nerve conduction velocity (NCV), and second, to determine if changes in NCV are dependent on age. We hypothesized that training would result in faster nerves in both young and older adults, albeit to a lesser extent in older adults. Forty-eight subjects (18-84 yrs) completed this study (n = 26 younger, 22 older). Motor NCV and maximal strength were recorded before and after 4-weeks of handgrip training in both limbs. Training was conducted 3×/week with the use of a grip training kit. Mixed-factorial ANOVAs revealed significant increases in NCV for both the young (*p* < 0.001) and older training groups (*p* < 0.001), but neither control groups (p > 0.05). The young training group showed increased handgrip strength after four weeks (*p* = 0.004), while the other groups did not. The results of this study suggest that resistance training may be a viable method to counteract age-related nerve deterioration. These outcomes have the potential to improve quality of life and generate greater independence for our older populations.

**NEW & NOTEWORTHY:** Aging is characterized by a slowing in nerve conduction speed, which could potentially affect how quickly an individual responds to unexpected incidents. Consequently, this slowing could increase the risk of falls and injuries. This study utilized resistance training as an intervention to mitigate age-related nerve deterioration. We show that motor nerve conduction velocity significantly increases in both young and older adults after 4-weeks of training.

## INTRODUCTION

Aging is a multidimensional process that is accompanied by several physiological and morphological changes which may affect one’s quality of life. It is well-established that declines in motor ability can be seen as early as the 4th decade (1, 2). Evidence supports the belief that functional ability, especially after the age of 60, is substantially reduced, and deterioration of the peripheral nervous system (PNS) may be one of the primary causes (3, 4, 5). Unfortunately, while there is substantial sarcopenia-related research focusing on preserving muscle mass and function, the collective research into counter-measures to protect peripheral nerve function is minimal.

Deterioration of the peripheral nerves is reflected by fewer functioning motor neuron axons and/or degradation of the motor axons. These outcomes result in the slower transmission of a nerve impulse supplying a target effector muscle, and can have functional consequences such as slower movements, reduced mobility or even trigger disordered movement, all of which may lead to a diminished quality of life (6, 7). Additionally, older adults exhibit varying degrees of loss in strength and muscle mass as a result of these effects and may become more susceptible to the development of sarcopenia (e.g., age-related reduction in muscle mass).

Commonly recognized in individuals with nerve injury or disease, declining PNS function often leads to neuromuscular dysfunction. However, age-related slowing of healthy motor nerves has been found in both humans and rodents (8, 9, 10). Previous research identifies a linear relationship between a slowing in nerve speed and age (≥ 60 yrs.) in those who are free of neuromuscular disease (11). While the support for an age-related slowing of nerves exists, there is much still to be understood regarding interventions that could facilitate positive adaptations within the peripheral nervous system.

Resistance training has long been prescribed to older adults as a means to long-term vitality. Individuals who remain active throughout their life have been known to have improved mobility, more independence, and greater life expectancy (12). Literature has shown that strength training may be a counteractive modality to nerve loss in previously injured nerves (13). However, it remains unclear if resistance training can alter nerve function in healthy non-injured adults. Moreover, resistance training has also been used in the prevention and management of sarcopenia (14, 15). While little is known about the effects of resistance training and nerve speed in healthy untrained individuals, there have been favorable findings in chronically trained athletes (16). Due to the benefits resistance training provides, it’s plausible to consider resistance training as a method to counteract losses in nerve function.

Although previous studies have investigated training and nerve speed in adults, few studies have focused on interventions that mitigate nerve speed loss and possible adaptations training may have. Additionally, our understanding of nerve function in untrained healthy individuals is still unclear. Further investigation is needed to determine the age-related changes in nerve speed function and the potential role exercise-based interventions may afford in resisting nerve deterioration. Therefore, the purpose of this study was 1.) to quantify the effects of resistance training on nerve conduction velocity, and 2.) to determine if age affects nerve plasticity in response to training. We hypothesized that resistance training would improve motor conduction velocity of the median nerve in both young and older adults in response to a hand-grip resistance training program. Second, we also hypothesized that the magnitude of adaptation was going to be significantly reduced in older adults.

## METHODS

### Participants

Forty eight participants (mean ± SD age = 43.9 ± 24.9) volunteered for this study (see Table 1 for descriptives). Participants completed an informed consent and health history questionnaire before beginning the study. All participants were apparently healthy and reported having no neuromuscular disease or neurological disorders. Further, all participants reported either having no upper body resistance training within the past six months or limited training (≤ 2 x per week). This study was approved by the University’s Institutional Review Board prior to data collection.

### Research Design

This study consisted of four groups, first separated by age based on our target ranges (young = 18-35 yrs old, and older = 60+ yrs old), and then pseudo-randomly assigned to either the intervention group or control group. This resulted in a young training group (YT, *n =* 14), young control group (YC, *n =* 12), older training group (OT, *n =* 14) and an older control group (OC, *n =* 8). Testing sessions were performed before (PRE) and after (POST) four weeks of the study. Motor nerve conduction and maximal hand grip strength testing were conducted during each testing session. Participants were asked to maintain normal daily activities throughout the study.

### Motor Nerve Function Assessments

Motor nerve conduction velocity (NCV) was collected using the incremental method. This quantifies NCV by obtaining the maximal m-wave response, also referred to as the compound muscle action potential (CMAP), at different segments along the median nerve and divides them by the distance between segments. The formula is shown in Figure 1.

**Figure 1.**
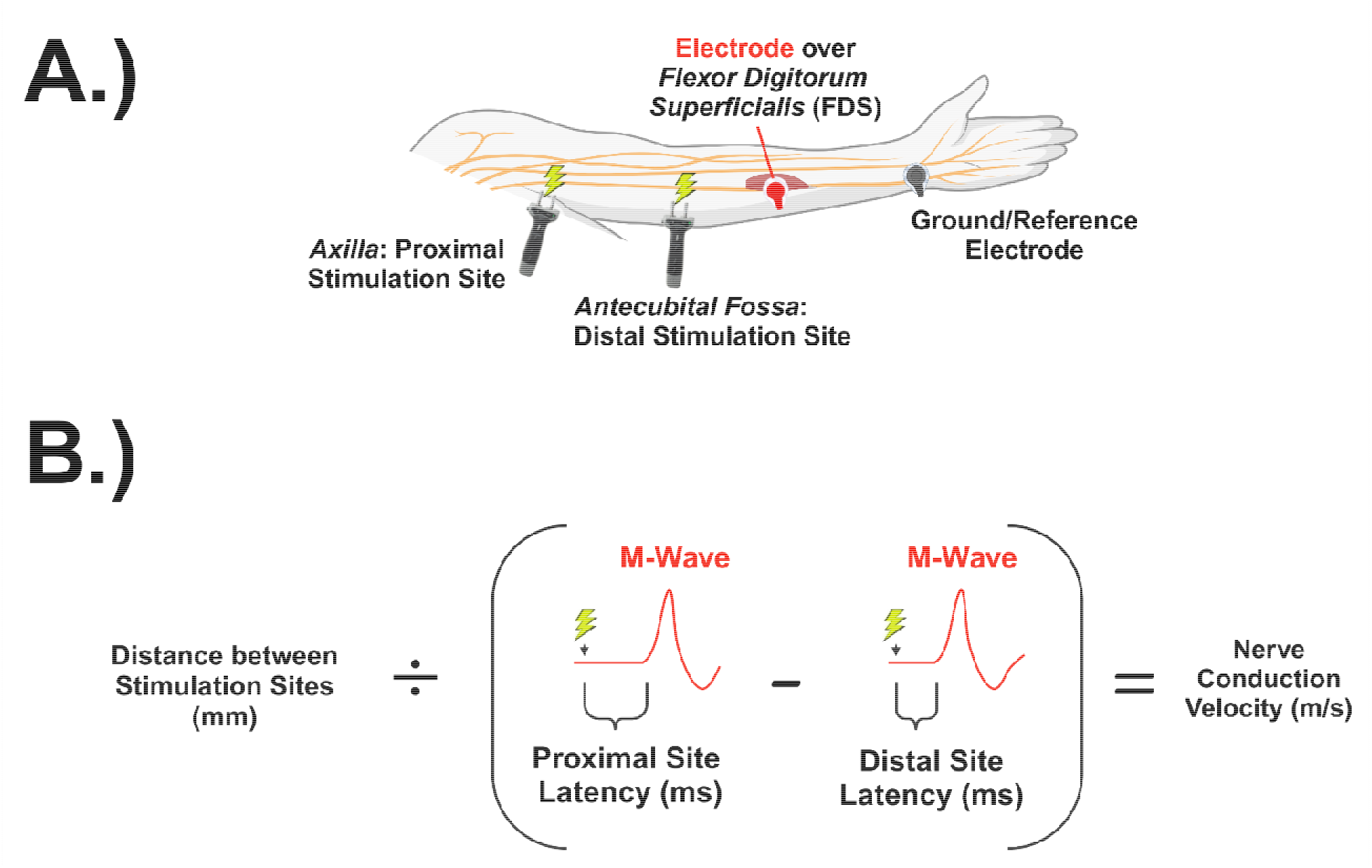
A.) A visual depiction of our nerve stimulation methodology. This shows the simulation sites (axilla and antecubital fossa) and electrode placement for a nerve conduction assessment of the median nerve. B.) A figure illustrating the calculation of nerve conduction velocity (m/s) from proximal latency (ms), distal latency (ms), and the distance between the stimulation sites (mm).

Using a clinical electrodiagnostic system (Cadwell Sierra Summit, Cadwell Industries, Inc., Kennewick, WA, USA), motor nerve conduction was assessed for the left and right median nerve. Disposable recording electrodes (20×27mm, Cadwell Industries Inc., Kennewick, WA, USA) were placed on the belly of the flexor digitorum superficialis muscle as verified by ultrasonography. The reference and the ground electrodes were placed on the tendon (flexor carpi radialis tendon) and the back of the hand (metacarpals), respectively. Prior to electrode placement, the skin over the specified sites were shaved, abraded, and cleaned with alcohol to ensure optimal signal quality.

Maximal m-waves were obtained at the axilla region and below the elbow of each arm. For assessment of the optimal stimulation site of the axilla region, the arm was palpated by having the participants flex their biceps. The optimal stimulation site for the elbow was located at the antecubital fossa near the biceps tendon. In a cathode-anode arrangement, a single stimulus (single square wave impulse) was applied in stepwise increments starting at 5mA. Each stimulus thereafter was increased by 5mA until no increases in m-wave amplitude were detected.

Following the assessment of maximal m-wave, NCV was obtained. A temperature probe was secured to the participant’s wrist to ensure consistent temperature (approx. 30°C) throughout testing (Med-Linket, Ltd, Shenzhen, Guangdong, China). A single supramaximal (e.g., 120% of maximal m-wave) stimulation was applied to each proximal and distal site (axilla and below the elbow) on both arms. The latency (m/s) of each stimulation was stored and recorded for later use in the determination of NCV. Additionally, the distance (mm) from the proximal stimulation site to the distal stimulation site was verified using a standard flexible tape measure. NCV was derived by the software included in the stimulation cart using the equation provided in Figure 1.

### Maximal Voluntary Contractions

Participants were asked to hold a hand dynamometer by their side and perform two maximal hand grip contractions per arm (Jamar, Sammons Preston Inc., Bolingbrook, IL, USA). Prior to the contractions, participants performed 2-3 warm-up contractions at half their maximal effort. Upon directions, participants were asked to raise the dynamometer to a 90° angle and contract maximally while exhaling each breath. Each contraction lasted 4-5 seconds with strong verbal encouragement given by the research team, and one minute of rest given between each trial. The highest value (kg) was considered the participant’s maximal hand grip strength (MVC).

### At-Home Resistance Training Intervention

Participants in the training groups performed bilateral hand grip resistance training three times per week for four weeks using a specialized hand-grip kit provided to them (NONJISPT, Shenzhen, Guangdong, China). Each kit contained an adjustable hand gripper, stress ball, grip ring, grip master, and finger stretcher resistance band. Additional grip rings (10-50 lbs; Refluxe, Shenzhen, Guangdong, China) and finger stretcher resistance bands (8-21 lbs; Portholic, Shenzhen, Guangdong, China) were used to induce a progressive overload in resistance for four weeks. Participants were provided with pictures and instructions for each exercise upon completion of the initial testing (PRE) session. They were asked to perform 12 training sessions (approximately 30-45 mins) over the 4 weeks according to their schedule. Each participant was asked to keep a training log to assist with accountability. The PI also conducted weekly check-ins (via phone call or text) with each subject to ensure compliance to the protocol and answer any potential questions. Subjects were excluded from the study if they noted three or more missed training sessions. Upon completion of the training, participants revisited the lab for a (POST) testing session and all previous measures were assessed. Those in the control group still received a hand-grip training kit after completion of the study, along with instructions on how to conduct the training.

### Statistical Analyses

All statistical analyses were performed using SPSS Version 26 (IBM, Armonk, NY, USA). Two four-way mixed factorial ANOVA model’s (age [young vs old] × group [training vs. control] × time [pre vs. post] × limb [left vs. right]) were conducted to determine changes in NCV and MVC strength. If warranted, dependent paired samples t-test were run. An alpha level of 0.05 was used for all comparisons.

## RESULTS

The results from the mixed factorial ANOVA showed no significant (age × group × time × limb) interaction for motor nerve conduction velocity (*p* = 0.937). Though, a significant group × time interaction was identified (*p* ≤ 0.001). None of the 2- or 3-way models that included age or limb as a factor were significant (p-values ranged from 0.216 – 0.987). Subsequent t-tests showed that both training groups had significant changes in NCV after training (YT: (t (13) = −5.733, (*p* < 0.001); OT: (t (13) = −4.694, (*p* < 0.001). Both training groups accounted for a 5.6% increase in NCV pre to post intervention. There were no significant findings for either control groups. The individual subject results are shown in Figure 2.

**Figure 2.**
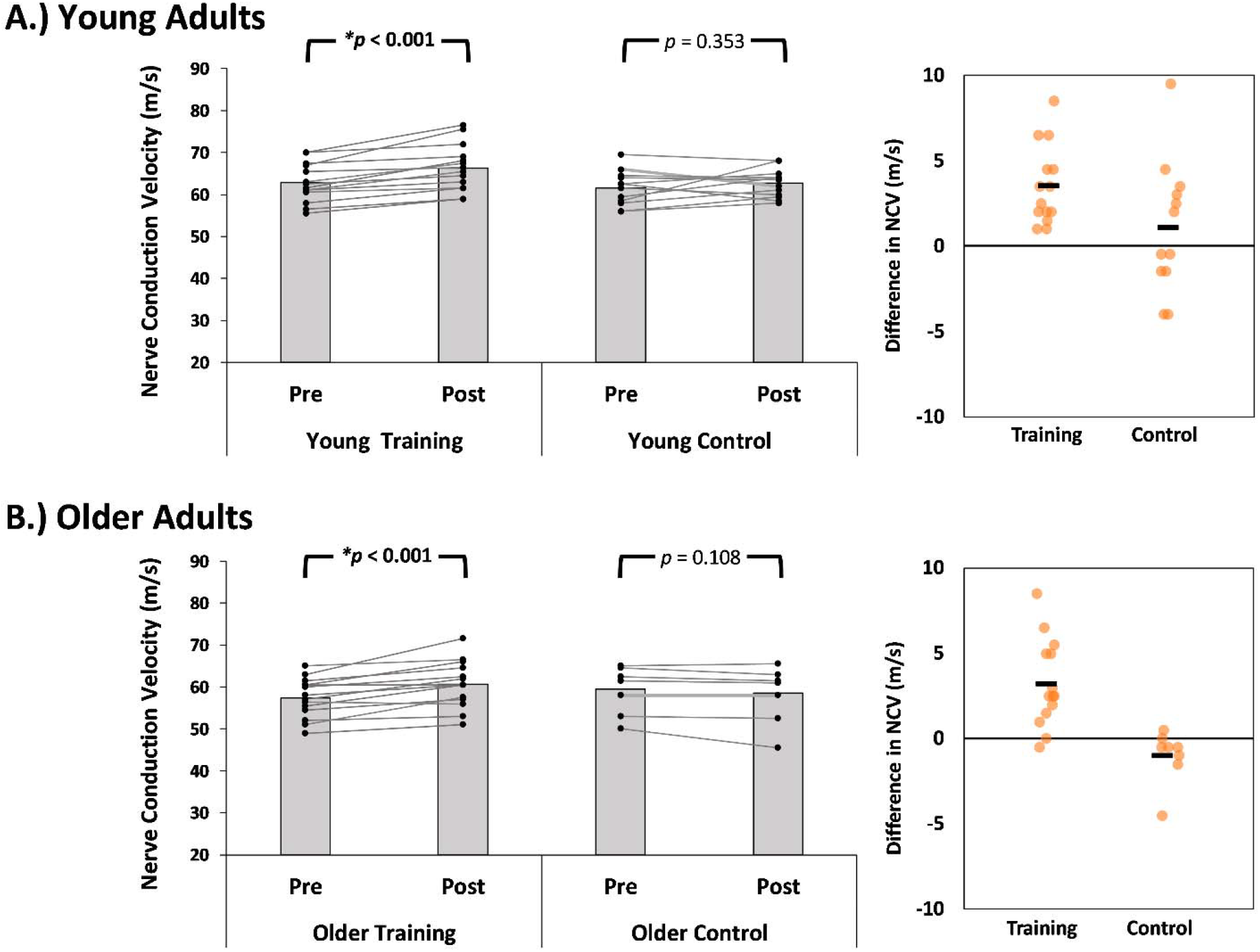
Results for nerve conduction velocity (NCV) before and after the 4-week training intervention for both the Young (panel A) and Older (panel B) adults. The gray bars represent group means, and the data points are individual subjects. Significant differences (*p* ≤ 0.05) are bolded and denoted (**\***). The right panel depicts individual change scores in NCV.

No significant (age × group × time × limb) interaction was observed for MVC strength (*p* = 0.970). However, significant interactions for (time × limb) and (group × time) (*p* = 0.018; *p* = 0.019, respectively) were found. The YT group demonstrated significant changes in MVC strength of both limbs pre to post intervention (left: (t (13) = −4.020, *p* = 0.001; right: (t (13) = −3.481, *p* = 0.004). A greater percentage of change was found on the left limb (8.9%) compared to the right (7.1%), which was expected as most individuals in this group were right handed (*n =* 12). No other significant changes were found among limbs or groups. The individual subject results are shown in Figure 3.

**Figure 3.**
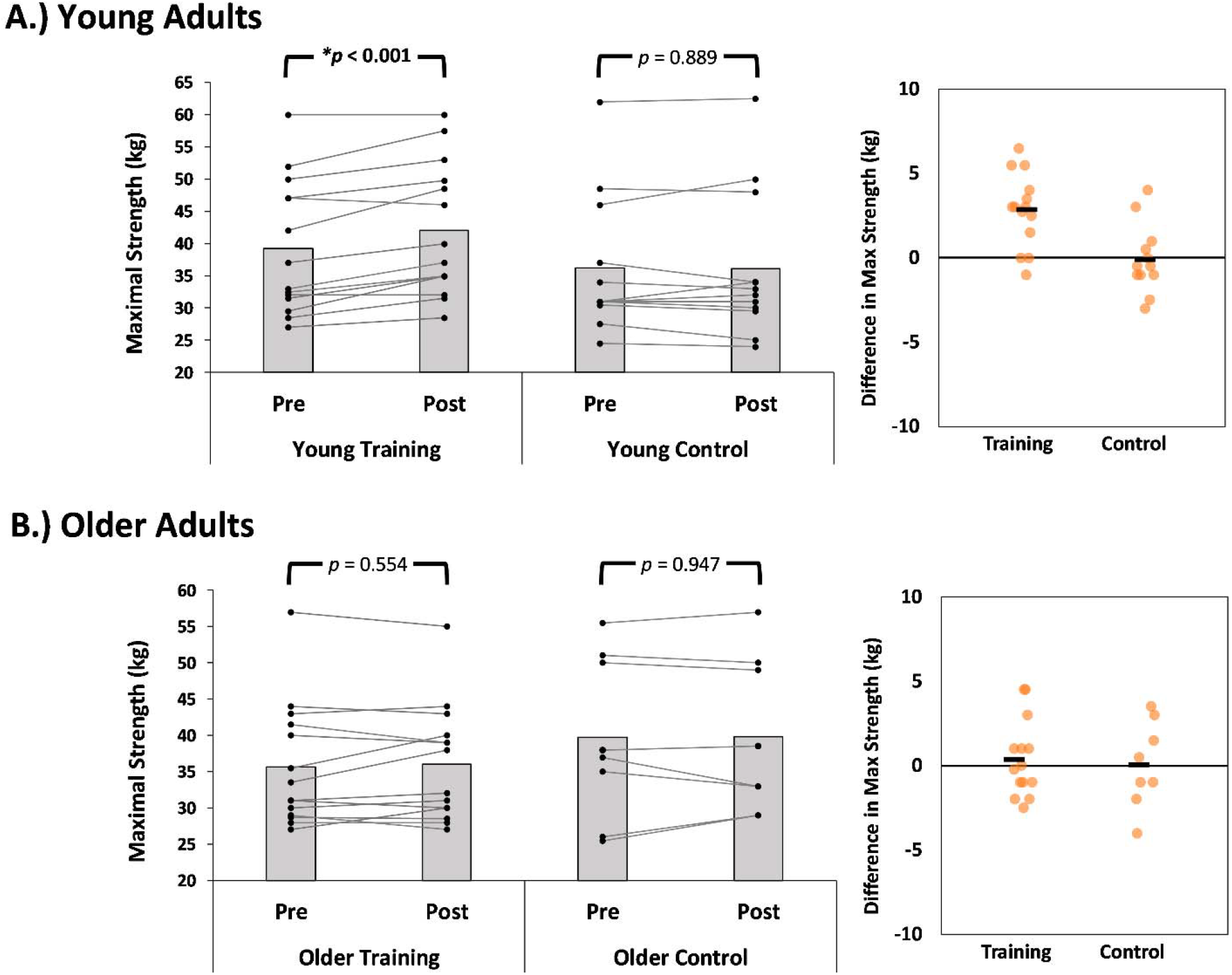
Results for maximal handgrip strength (MVC) before and after the 4-week training intervention for both the Young (panel A) and Older (panel B) adults. The gray bars represent group means, and the data points are individual subjects. Significant differences (*p* ≤ 0.05) are bolded and denoted (**\***). The right panel depicts individual change scores in MVC.

## DISCUSSION

This study aimed to examine the trainability of peripheral nerve function as well as its dependency on age. We found that 4-weeks of hand-grip strength training increased both strength and median nerve conduction velocity in both forearms. The changes in strength in the young group affirm the efficacy of the strength training intervention. Although we hypothesized that training would lead to improvements in nerve function, we did not expect the large magnitude of adaptations demonstrated in this study from such a short intervention. The 4-week duration of the intervention was chosen more for project feasibility than it was for it representing any sort of optimal duration. As such, we expected smaller and less robust gains in which there would be some non-responders present. The lack of significant improvements in strength in the older group can likely be attributed to the intervention duration. However, the development that every participant in the 2 intervention groups showed improvement in nerve conduction velocity was unexpected, and requires follow-up studies that examine potential underlying mechanisms to better understand this. A few studies have investigated various exercise prescriptions and their subsequent effects on conduction velocity. In a study on strength and sprint training, Sleivert et al. (17) found that conduction velocity significantly increased after 14 weeks in untrained young males. However, findings on chronically trained (e.g., weightlifters and athletes) individuals show varied results (18, 19). However, these studies were only in young males and young male athletes, and did not include women or older adults.

### Adaptations Dependency on Age

Unexpectedly, older adults had just as strong of an adaptation to the training as the young adults. We anticipated that the magnitude of change would be significantly reduced compared to younger participants (i.e. reduced plasticity). For example, previous literature has found attenuated responses in measures of muscle size (20, 21) and muscle strength (22, 23) after training in older adults. Based on these findings, we incorrectly anticipated a similar pattern in nerve function would occur.

### Adaptations Dependency on Limb

It was also slightly surprising that limb was not a factor in any of the statistical models. We expected the dominant limb to be slightly more trained than the non-dominant, and as a result, hypothesized the dominant limb would have reduced gains. It has been reported that the dominant limb is often stronger and more trained (24, 25). However, a study by Coombs et al. (26), showed that three weeks of either right or left-handed strength training demonstrated symmetrical interlimb (“cross-education”) effects between dominant and non-dominant wrist extensors. In contrast, Farthing et al. (27) found that muscular strength was unidirectional in right hand dominant individuals after six weeks of unilateral training. It is suggested that the characteristics of a task and the training paradigm may influence outcomes, which could explain limb being a non-factor in our present study.

### Potential Mechanisms

The findings shown in the present study lead to a particularly intriguing follow-up question; what are the potential mechanisms underlying this improvement in nerve conduction velocity? It has been well established for nearly a century that there is a strong positive relationship between the diameter of an axon and its conduction velocity (28, 29). Therefore, one possible mechanism for the neurons to get faster is for them to also get larger (wider axonal diameter). There is substantial research into the training-induced hypertrophy of skeletal muscle fibers, but it is relatively unknown as to whether neurons are also capable of hypertrophy in response to training. The opposing result of nerve atrophy has been investigated in-depth, and does demonstrate a continued marriage between axonal size and conduction velocity (i.e. as one goes down, so does the other)(30). Therefore, although speculative, it is at least plausible that the 2 variables may continue to be paired for improved adaptations as well. Future studies examining strength training interventions should include both a measure of nerve conduction velocity as well as a measure of axonal size (or at least an estimate, such as nerve size). Another possibility for the nerve includes potential changes in axonal excitability (31). However, since our measure of NCV requires a compound muscle action potential (M-wave), downstream mechanisms, such as increased efficiency of transmission across the neuromuscular junction or faster conduction velocity in the muscle fibers, cannot be ruled out.

### Limitations and Future Directions

While our study presents new and exciting findings, there are some conceivable limitations to this study design. First, the level to which our participants were “untrained.” While inclusion criteria allowed participants to exercise (≤ 2 x per week), this may have contributed to our findings. Several older adults reported being active, and a few participated in racket sports, which may have influenced the changes observed. Second, it is possible that compliance may have been a factor. While participants were asked to track their training, one can never entirely ensure participant compliance with the training program. Thirdly, it is possible that our intervention was too short (only 4 weeks), and future studies with longer intervention durations may provide stronger evidence as to the plasticity of nerve conduction.

More thorough investigations are needed to determine the underlying mechanisms of this study’s findings. Additional research should explore the functional properties of the spinal and supraspinal systems and their contributions to peripheral nerve transmission. Moreover, it is important to identify potential interventions and optimal prescriptions that may support the neural consequences of aging.

## CONCLUSIONS

These findings support our hypothesis that hand grip training improves motor conduction velocity in young and older adults. This suggests that resistance training may be a robust method to counteract NCV deficits in the short term, although more research is still needed. The results of this study could aid clinicians in exercise prescription for individuals needing to improve nerve conduction and motor function. The significance of this research line has the potential to improve the quality of life and generate greater independence for our older populations.

## Data Availability

All data produced in the present study are available upon reasonable request to the authors

## ACKNOWLEDGEMENTS

None.

## AUTHOR CONTRIBUTIONS

J.E.S. conceived and designed research; C.M.S., S.M.R., M.L.D.S., and J.E.S., performed experiments; C.M.S., M.P., and J.E.S., analyzed data. J.M.D., and J.E.S., interpreted results of experiments; J.M.D. and J.E.S., prepared figures; J.M.D. and J.E.S., drafted manuscript; C.M.S., S.M.R., M.L.D.S., M.P., J.M.D., and J.E.S., edited and revised manuscript; C.M.S., S.M.R., M.L.D.S., M.P., J.M.D., and J.E.S., approved final version of manuscript.

